# Video-Audio Neural Network Ensemble For Comprehensive Screening Of Autism Spectrum Disorder in Young Children

**DOI:** 10.1101/2023.06.28.23291938

**Authors:** Shreyasvi Natraj, Nada Kojovic, Thomas Maillart, Marie Schaer

## Abstract

A timely diagnosis of autism is paramount to allow early therapeutic intervention in preschoolers. Deep Learning (DL) tools have been increasingly used to identify specific autistic symptoms, and offer promises for automated detection of autism at an early age. Here, we leverage a multi-modal approach by combining two neural networks trained on video and audio features of semi-standardized social interactions in a sample of 160 children aged 1 to 5 years old. Our ensemble model performs with an accuracy of 82.5% (F1 score: 0.816, Precision: 0.775, Recall: 0.861) for ASD screening. Additional combinations of our model were developed to achieve higher specificity (92.5%, i.e., few false negatives) or sensitivity (90%, i.e. few false positives). Finally, we found a relationship between the neural network modalities and specific audio versus video ASD characteristics, bringing evidence that our neural network implementation was effective in taking into account different features that are currently standardized under the gold standard ASD assessment.

## Introduction

Autism spectrum disorders (ASD) are a group of neurodevelopmental disorders characterized by a wide range of symptoms across two main dimensions: social and nonsocial. The social dimension spans the domains of verbal and nonverbal social communication, interpersonal interactions, and socio-emotional reciprocity. The symptoms in the nonsocial dimension include the repetitive and restricted patterns of behaviors and sensory issues exerting various levels of impairment in the everyday social functioning and autonomy of the individual^1^. ASD can be present with or without cognitive and language delay, widening the breadth of clinical presentations. In absence of any reliable biomarker identified to date, the ASD diagnosis depends on precisely identifying entangled behavioral patterns. The current gold-standard diagnostic procedure is the product of convergent clinical and research work over the past four decades aimed at standardizing the clinical practice of accurately detecting and measuring symptoms^2–5^. The heterogeneity of ASD manifestations is a major obstacle to timely diagnosis and personalized medicine in autism^6^. Despite the diagnosis now being possible at ages younger than 2, the latency between the first observable signs of autism and the age of diagnosis usually spans years^7^. Convergent findings established the efficiency of early and intensive behavioral intervention in improving outcomes of individuals with autism^8–10^. According to current clinical guidelines, early diagnosis is a crucial catalyst for early intervention^11^, which in turn is found to be most efficient when applied during the period of enhanced brain plasticity^12^. As such, an efficient and reliable screening is a critical first step to provide access to individualized and adequate support for the children who might need it.

Machine learning and computer vision models have been increasingly used to implement objective and reliable detection of autism to eventually overcome the shortcomings of the classical questionnaire-based screening tools^13,14^. The first computer vision (CV) models to detect autism focused mostly on nonsocial, motor symptoms^15^. These models have the advantage of being relatively context-independent and thus, they are easier to operationalize than the symptoms in the social domain with a more contextual ontology. Social symptoms often transcend the isolated instances of behaviors and arise from specific mismatches in the dynamic between agents and their social context. In the search for potential biomarkers of ASD disorder, previous studies investigated behaviors such as postural asymmetry^16^, gait patterns^17^, gesture motion energy^18^, or frequency of stereotyped behaviors^19,20^. These results have validated the possibility of automating the measurement of known behavioral features in autism. For social symptoms, the development of automatized measures has often implied standardization of the social conditions. For example, in the response to name paradigm, the child was engaged on a tablet screen while her/his name was being called^21^. Given the complexity of social symptoms, over-standardization seems a necessary first step to allow a precise readout by carefully controlling the feature context at the expense of less ecological feature presentation. Unsurprisingly, there have been fewer attempts to automatize behavior measures of autism in the context of free social interactions. Using segments of free-play interactions, Budman and colleagues^22^ measured instances of approach/avoidance behaviors in children with ASD. Similarly, a laboratory task measuring the dyadic synchrony^23^ highlighted a specific “synchrony signature” in dyads involving at least one individual with ASD. The finding of reduced interpersonal synchrony was corroborated by an observational study using a more ecological approach of semi-standardized interactions^24^. In previous work^25^, we developed a machine learning approach that considers behaviors in their natural (unconstrained) context of social interaction with an adult during semi-structured diagnostic assessment (ADOS)^4^. The focus was on nonverbal social interaction features through pose estimation^26^. The classifier is sensitive to temporal social interaction features in a low dimension domain and distinguishes between ASD and TD children with an accuracy of 80%^25^. Thus, the unconstrained, dynamic, nonverbal features of social interaction hold promise for developing novel automated screening tools for ASD diagnosis.

The above-mentioned studies focused on social interaction analyses from a computer vision perspective, considering mostly visual aspects of the social exchange. A line of research delved into another dimension of information that can be analyzed with automatized measures, namely vocal and verbal features. Indeed, atypicalities in vocal and verbal production were noted in the first descriptions of the disorder^27,28^ and still are considered one of the core diagnostic features of the disorder^1^. Individuals with ASD often present language delay, which, historically, has constituted one of the principal defining features of the disorder^29^. Longitudinal studies have shown that, throughout their development, around 80% of autistic individuals develop functional language. Nevertheless, atypicalities in acoustic language features, particularly prosody, remain a salient feature of their verbal production. Indeed, alteration of specific acoustic features of vocal productions, such as pitch mean and variability, have been consistently reported in autistic individuals of all ages^30,31^. As altered prosody can also be measured without phrase speech, measuring specific acoustic vocal features could become a reliable marker of autism, even early on: Santos et al.^32^ have achieved an impressive accuracy of 97% in detecting autism at only 18 months old. Most studies use either manual diarization protocols or voice extraction fostered by using a child-worn device (such as LENA^33^), to analyze the characteristics of the child’s voice. The tedious process of manual voice identification, or the need to use a specific child-worn device, limits the possibility of analyzing video recordings that would have been collected without the specific device in diverse clinical or research contexts, thus restraining scalability. Also, focusing only on the child’s vocal productions alone, without analyzing them in the context of social interaction, inherently reduces the richness of the information that can be extracted. Several studies have shown the promising potential of analyzing reciprocal features of the conversation, such as turn-taking^34,35^. Finally, prosodic features of the adult interacting with the child have also been shown to be very informative, as caregivers or psychologists tend to attune their prosody to the child that they are addressing and align the complexity of their verbal production to the child’s language structure^36,37^. As such, one could hypothesize that the entire audio soundtrack of a social interaction contains a rich set of information about both the characteristics of the child’s vocal production as well as a signature of the mutual vocal & verbal choreography that has the potential to represent a powerful autism screening tool. To our knowledge, no study has tried to use a global acoustic pattern of social interaction (without explicit separation of speaker channels) for autism diagnosis prediction.

The present study builds upon our previous work that focused on the nonverbal features of social interaction to identify preschoolers with autism^25^. Here we assess the possibility of yielding an accurate classification of autism spectrum disorder based on the acoustic information from the semi-structured social interaction between a child and an adult. We adopt a comprehensive strategy for the audio analysis as in our video analyses. We consider the entire audio band of the social interaction session recorded in the context of the diagnosis session (Autism Diagnosis Observation Schedule-ADOS^3,4^). We preserve the natural flow of social interaction acoustics features and test if this global acoustic pattern is predictive of ASD diagnosis. We then test the prediction accuracy achieved by video and audio neural networks alone could be improved if the results of the two networks were combined. Finally, we analyzed the relationship between the audio and video neural networks concerning measures of autistic symptoms to cross-validate their clinical relevance. In the present study, we used a sample of 160 participants (80 typically developing (TD) children and 80 children with ASD, aged from 1.1–5.1 years old) who had undergone a video-recorded ADOS assessment. Our video neural network distinguished children with ASD with an accuracy of 80%, while the accuracy of the audio network was 79%. The combination of both networks yielded a prediction accuracy of 83%.

## Results

### Autism Prediction Using Video Features

To predict autism diagnosis solely from ADOS video features, we used a slightly modified approach compared to the one used in our previous study^25^ (see Methods). We used pose estimation on the ADOS recordings and subsequently divided them into 5-second segments, then implemented a modified version of the Convolutional Network VGG16^38^ Long Short-Term Memory (LSTM) recurrent neural network architecture^39^ (see Figure 1). We then trained the neural network using an 80-20 validation split.

**Figure 1.**
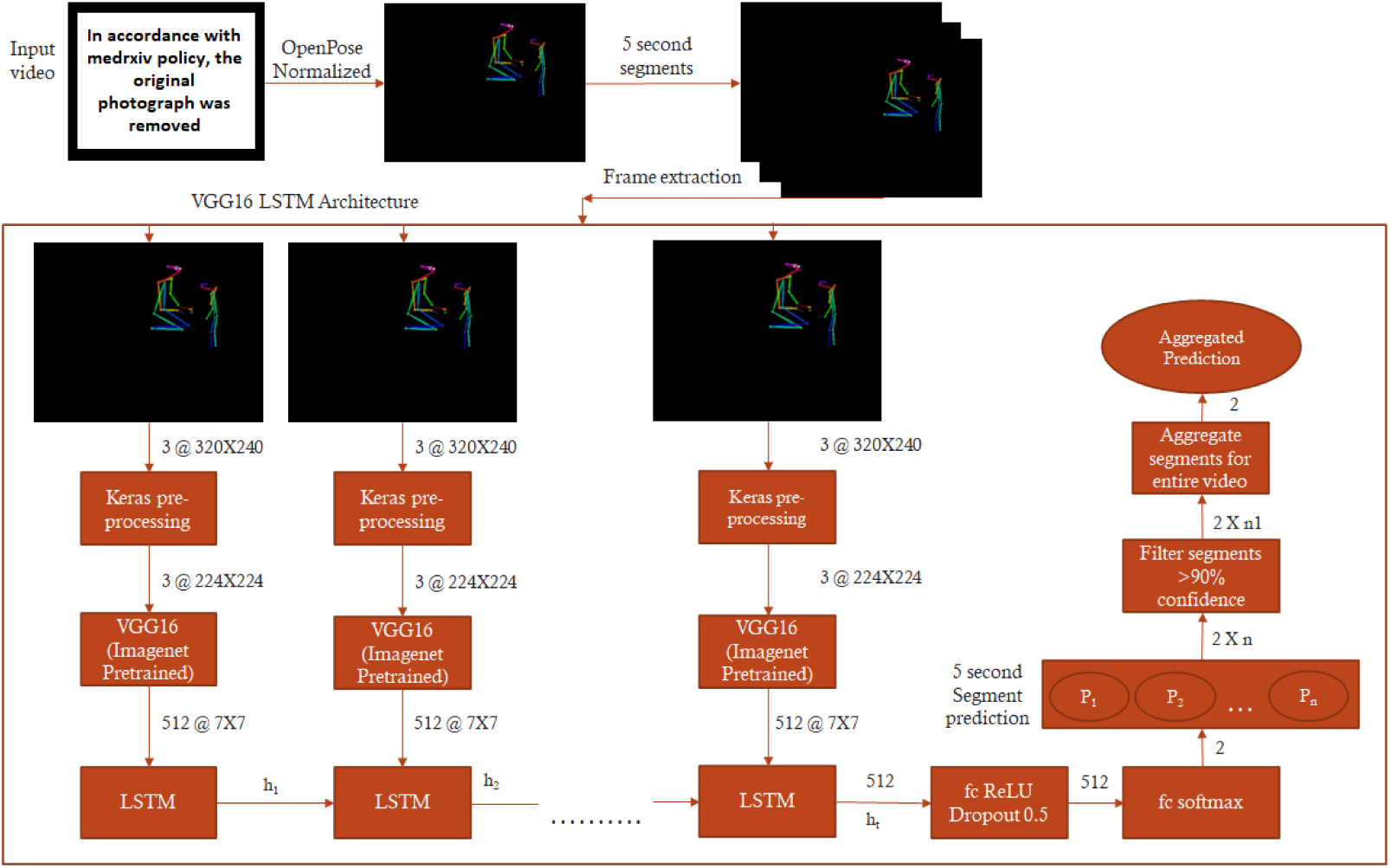
The Imagenet pre-trained Convolutional Network VGG16 is used to extract high dimensional features from 5-second segment frames and fed into a Long Short-Term Memory (LSTM) network operating with 512 units. Finally, the output of the LSTM is followed by 512 fully connected ReLU-activated layers and a softmax layer yielding two prediction outputs for the given segment. The segment-wise classification is aggregated for the video’s duration to obtain a final prediction value (ranging from 0 to 1) that we denote as “ASD probability.”. The entire video is classified as belonging to a child with ASD if the mean value of ASD probability is superior to 0.5.

Our model achieved an 80% prediction accuracy and an F1 score of 0.789 (see Table 1), confirming previous results using a smaller dataset^25^.

**Table 1.** Confusion Matrix representing the accuracy, precision, recall, specificity, negative predicted values, and F1-score for Audio, Video, and Ensemble Neural Network Models on a testing set of 80 videos. We also report on two other models for which we take the final prediction as ASD if either audio or video model predicts a subject as ASD (*OR Model*) and we take the final prediction as ASD only if both models predict a subject as ASD (*AND model*).

| Parameter | Video Model | Audio Model | Ensemble Model | OR Model | AND Model |
| --- | --- | --- | --- | --- | --- |
| Accuracy | 0.8 | 0.788 | 0.825 | 0.825 | 0.763 |
| Precision (Positive Predicted Values) | 0.833 | 0.811 | 0.861 | 0.783 | 0.889 |
| Recall (Sensitivity) | 0.75 | 0.75 | 0.775 | 0.9 | 0.6 |
| Specificity | 0.85 | 0.825 | 0.875 | 0.75 | 0.925 |
| Negative Predicted Values | 0.773 | 0.767 | 0.796 | 0.882 | 0.698 |
| F1 Score | 0.789 | 0.779 | 0.816 | 0.837 | 0.716 |

### Autism Prediction using Audio Features

We then explored the potential of accurately predicting autism from the full audio band of the ADOS assessment. After normalizing the audio recordings and splitting them into 10-second segments, we extracted several acoustic features^40–44^ that were then passed through a convolutional neural network (see Figure 2 and Methods). Following standard model hyperparameter tuning procedure, we deployed the 80-20 training validation split. This model yielded a final accuracy of 78.8% (F1 score: 0.779) in identifying autism solely on the audio track of the ADOS assessment (see Table 1).

**Figure 2.**
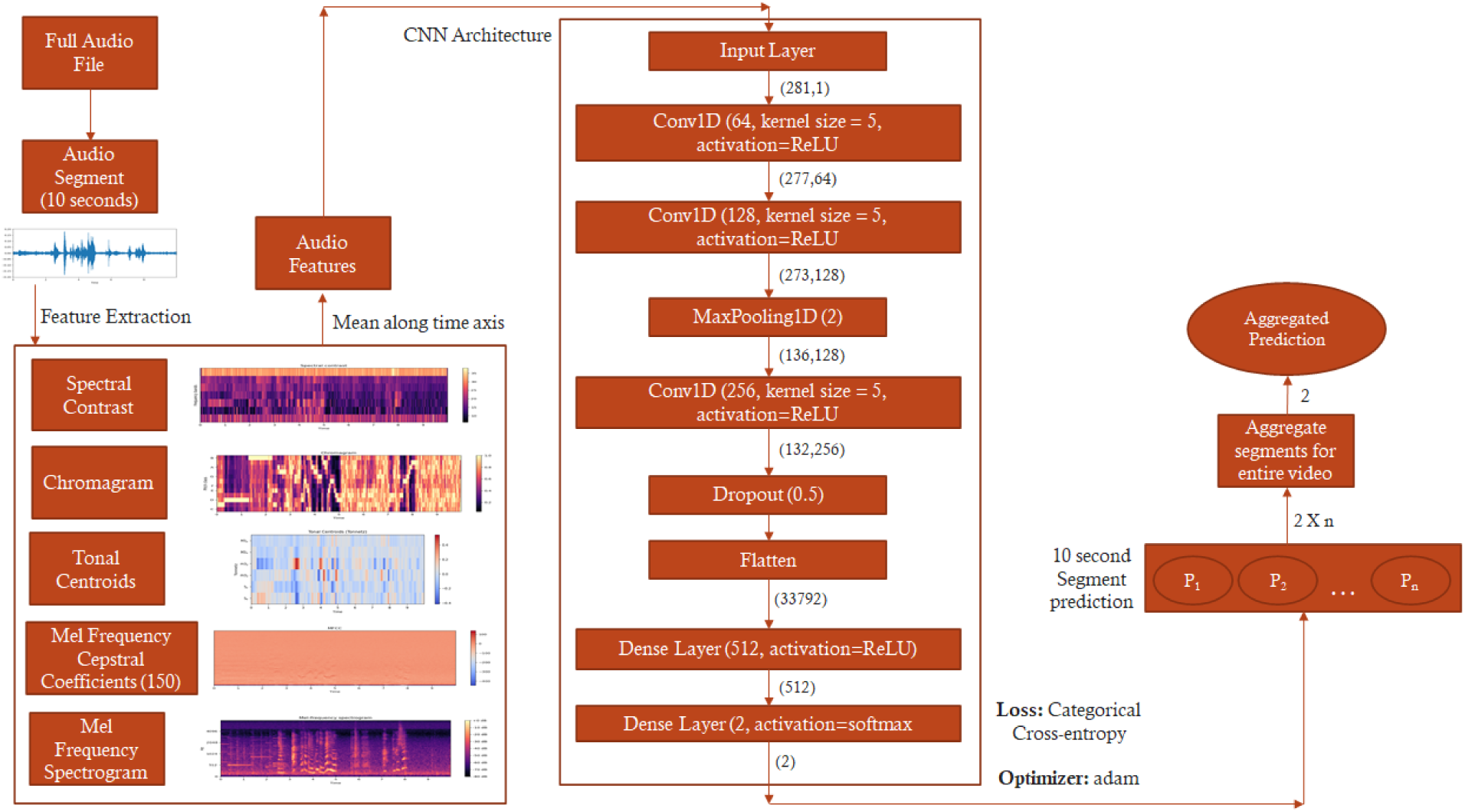
Audio neural network architecture. We used Librosa to extract Mel Frequency Cepstral coefficients, Mel Spectrogram, Tonal Centroid, Chromagram, and Spectral Contrast features. The neural network takes mean along the time axis for these features and inputs them into a convolutional neural network consisting of two one-dimensional convolutional layers. The segment-wise classifications are aggregated for the duration of the audio to obtain a final prediction value (ranging from 0 to 1), which refers to ASD probability. The entire audio band is classified as belonging to a child with ASD if the mean value of ASD probability is superior to 0.5.

### Autism Prediction using Combined Video and Audio Modalities

We explored the potential of combining the two modalities (Audio & Video) to tune the properties of our final prediction model regarding sensitivity, specificity, or accuracy (see Table 1). The *OR Model* yielded the highest sensitivity (0.9), with the lowest specificity (0.75). Such a model with a low rate of false negative results could prove useful as a screening tool, especially provided its high accuracy (82.5%) (see Supplementary Figure S4 A). Conversely, the *AND Model* gave a low rate of false positives, with the highest specificity (0.925), lowest sensitivity (0.6) and low accuracy (76.3%) (see Supplementary Figure S4 B). The best trade-off between the sensitivity and specificity was obtained using a decision tree (see Figure 3), where we retained the prediction from the video-neural network when the confidence was higher than 60%, otherwise replaced by the prediction from the audio neural network. This approach yielded an accuracy of 82.5% over the 80 videos from the testing sample (see Table 1). In only 8.75% of the cases, both neural networks failed to correctly predict autism spectrum disorder in children (see Supplementary Figure S5).

**Figure 3.**
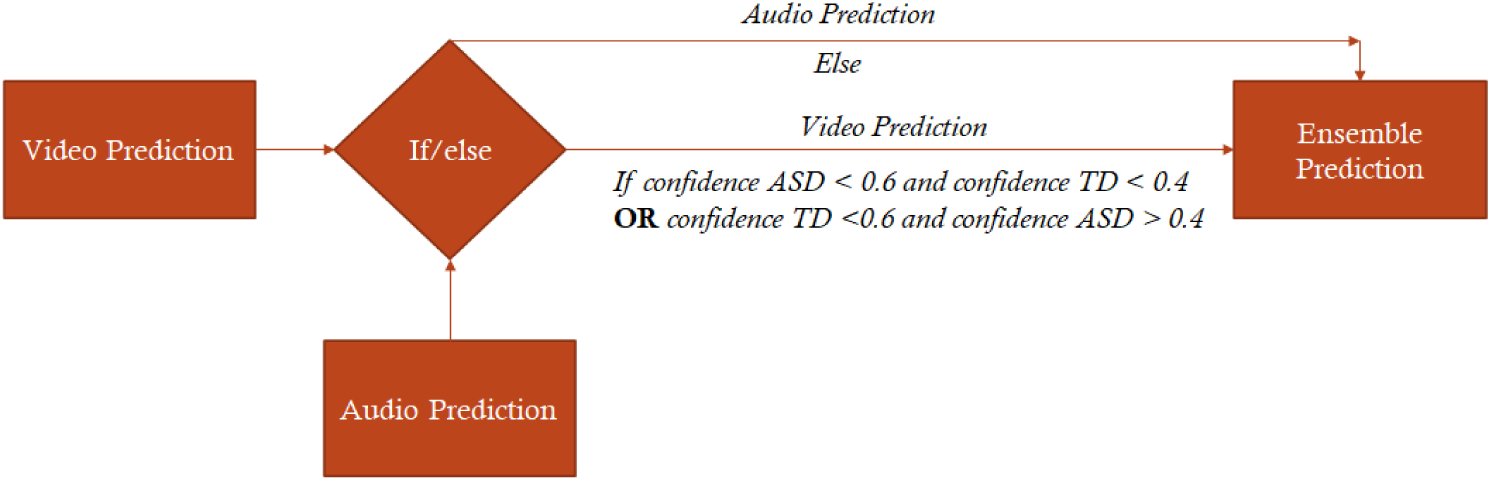
Decision tree combining predictions from combined video and audio neural networks. If the confidence of the video neural network is less than 60%, the model uses predictions provided by the audio neural network.

### Audio and Video Neural Network derived ASD Probability in the light of the Clinical Phenotype

Relating the accuracy of the neural network with specific ASD characteristics associated with better discrimination audio and video modalities, we used a random forest regression model^45^ for each modality to estimate the contribution of each of the 27 ADOS items to the confidence of the ASD prediction. For the video neural network, we found that the ADOS items that were more strongly associated with the ASD probability were the Initiation of Joint Attention, Quality of Social Overtures, the presence of Unusual Eye Contact as well as the occurrence of Pointing Gestures (see Figure 4 A & B). For the audio neural network, the ADOS items that were most strongly related to the ASD probability were the occurrence of Showing Behavior (to share interest), Pointing Gesture, Unusual Sensory Exploration, and Hand Mannerisms (see Figure 4 C & D).

Finally, we created a third random forest model to normalize and compare the Increased Mean Square Error obtained from the audio and video neural network for the 27 items of ADOS. These normalized values were then used to create a regression model^46^ to segregate ADOS items that were dependent on video, audio, or both the neural network’s confidence (see Figure 5). Our results show that the 3 most discriminant items based on video predictions were Initiating joint attention (IJA), Quality of social overtures (SO), and Visual contact. For the audio predictions, the 3 most discriminant items were Sensory interests, Showing, and Hand mannerisms. The least consistent items across audio and video predictions were Showing, Initiating Joint Attention (IJA), Giving, and Gestures. Strikingly, a large body of the ADOS items was consistent across both audio and video predictions (i.e., close to the regression line in Figure 5). Indeed, with a few exceptions, all ADOS-derived measures of symptoms are multimodal and reflect a cumulative (for the session duration) appreciation of behavioral patterns. This result is promising as it shows that our neural network implementation effectively considered different features currently standardized under the gold standard ASD assessment.

**Figure 4.**
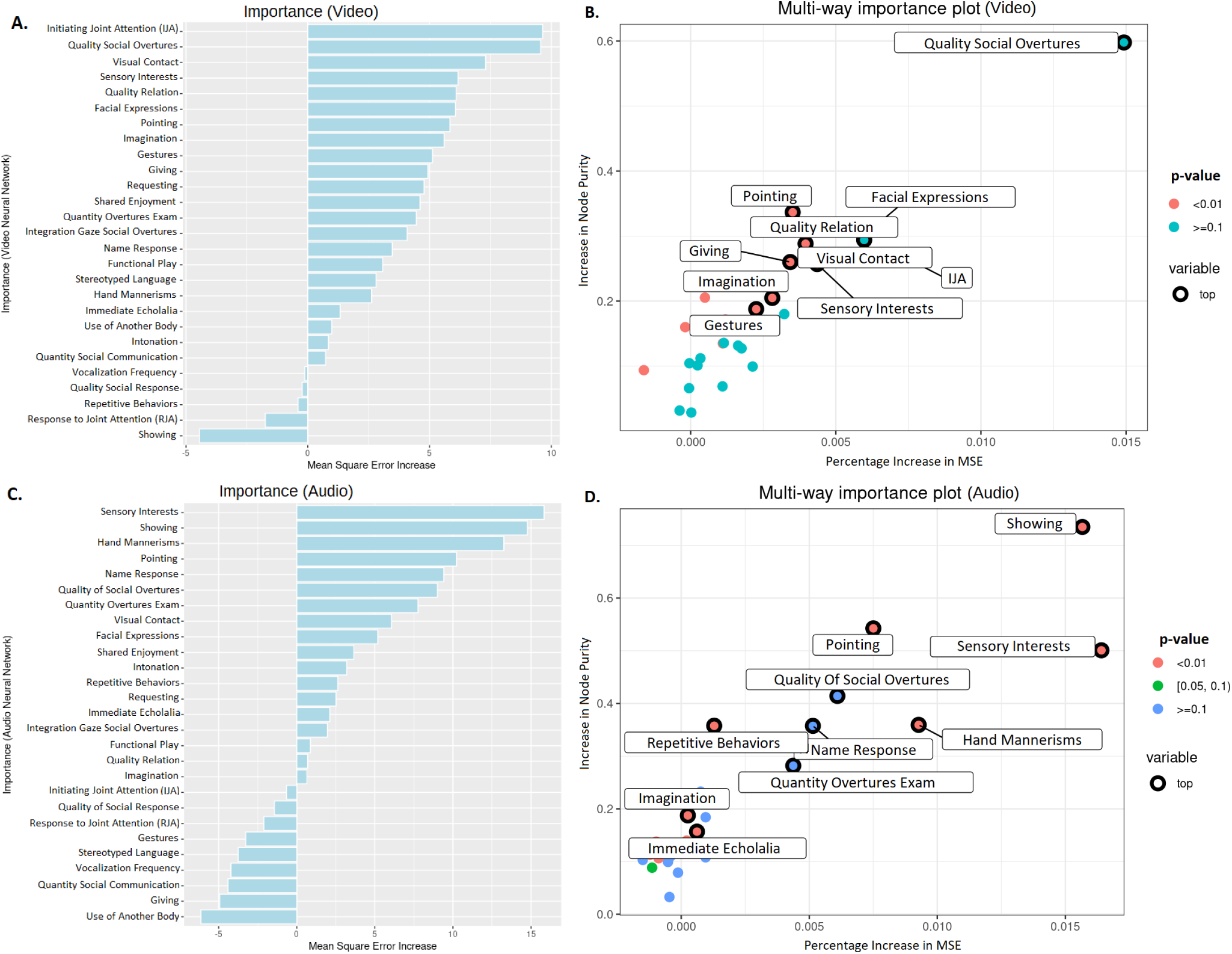
**A**. Sorted values of increase in Mean square error (%IncMSE) of the 27 ADOS-2 items against the confidence of the audio neural network predictions. **B**. A multi-way importance plot highlighting the 10 most important ADOS-2 items for estimating the confidence of the audio neural network via random regression forest modeling. **C**. Sorted values of increase in MSE for the 27 ADOS-2 items against the confidence of video neural network predictions. **D**. A multi-way plot highlighting the 10 most important ADOS-2 items for estimating the confidence of the video neural network via random regression forest modeling.

**Figure 5.**
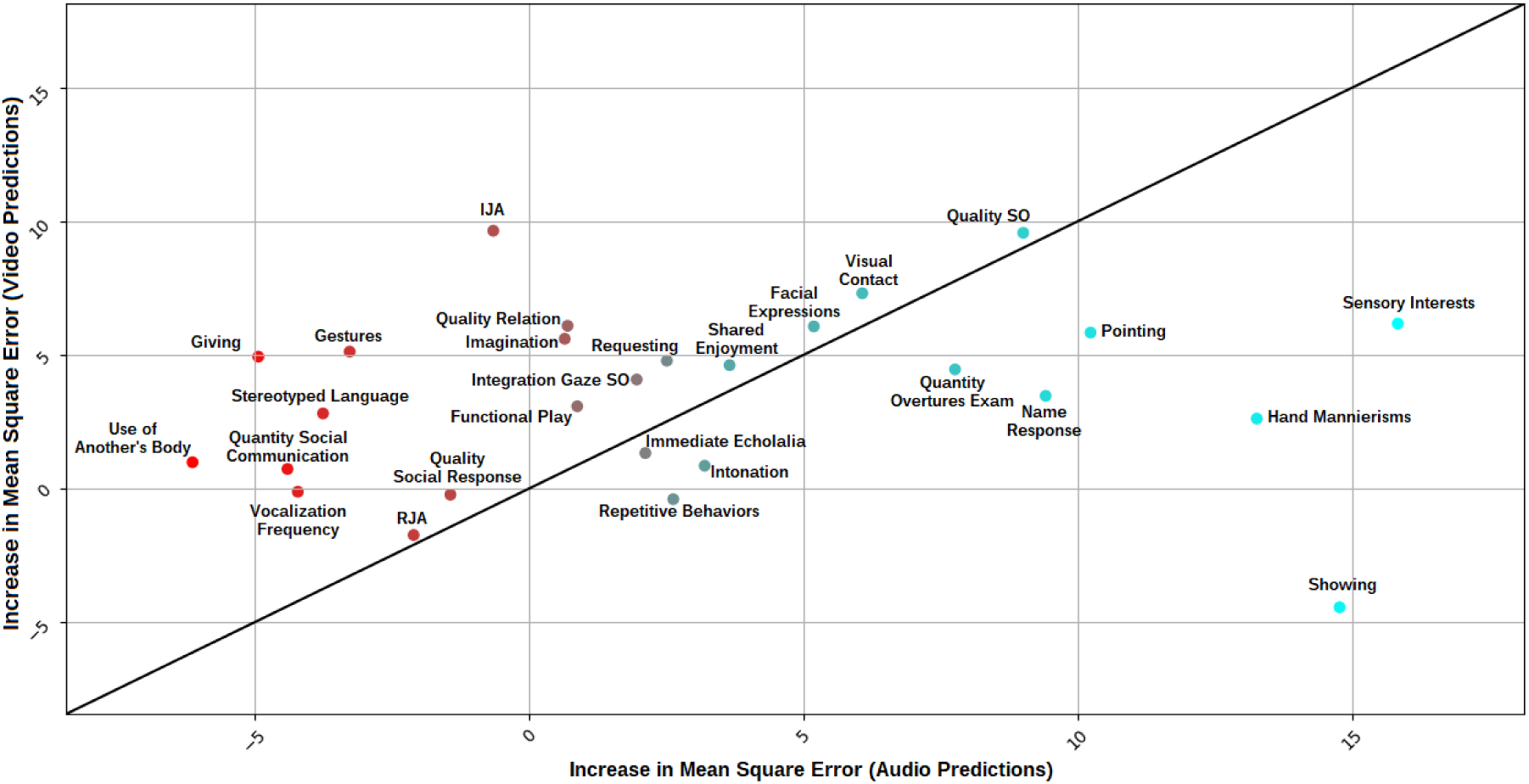
Residuals obtained after running the regression model over the 27 ADOS-2 items and computing the Increased Mean Square Error with respect to them predicting the confidence of the video or audio neural network in the random forest. The ADOS items included were: Frequency of Spontaneous Vocalization, Intonation of Vocalizations/Verbalizations, Immediate Echolalia, Stereotyped/Idiosyncratic Use of Words or Phrases, Pointing, Gestures, Unusual Eye Contact, Facial Expressions Directed at Others, Integration of Gaze and Other Behaviors during Social Overtures (SO), Shared Enjoyment in Interaction, Response to Name, Requesting, Showing, Giving, Spontaneous Initiation of Joint Attention (IJA), Response to Joint Attention (RJA), Quality of Social Overtures, Amount of Social Overtures/Maintenance of Attention: Examiner, Quality of Social Response, Quantity of Social Communication, Overall Quality of Rapport/Relation, Functional Play with Objects, Imagination / Creativity, Unusual Sensory Interest in Play Material/Person, Hand and Finger, and Other Complex Mannerisms, Unusually Repetitive Interests or Stereotyped Behaviors. The most discriminant features for video neural network are IJA, Quality of Social Overtures, and visual contact. Moreover, Sensory Interests, Showing, and Hand mannerisms are the 3 most discriminant features for the audio neural network.

## Discussion

We presented a multi-modal approach to the automated detection of autism based on semi-structured interaction between a child and an examiner. The present work extends our previous work on the automated identification of autism in young children using spatiotemporal features from the pose estimation applied to videos of semi-structured social interaction^25^. Here, using the late fusion approach^47^, we combine the results from the neural network using the pose estimation with the model operating on the acoustic features of the same interaction scenes used for training the video neural network. Relying on only visual information, we obtain an accuracy of 80% over a larger sample and further strengthen the evidence that autism diagnosis can be robustly identified from pose estimation in preschoolers. Using solely the audio information, we obtained a similarly high prediction accuracy (78.8%). While combining the video and audio models did not significantly improve the prediction accuracy, we found that different combinations of the predictions from the neural networks could result in models with either higher sensitivity (0.9) or higher specificity (0.925), bringing flexibility for adapting screening models depending on the needs. For further validation, we examined which ADOS items better discriminate autism diagnosis depending on the modality used for automated classification. We found that the quality of eye contact, as well as social overtures, are better captured in the visual neural network. Sensory interests and mannerisms were better captured in the audio neural network

Our results are intriguing because using only acoustic features or combining them with video features yields a high accuracy (i.e., ≈80%), comparable to video features only. The high accuracy of the audio neural network is particularly striking because the audio recordings used for classification contained the full, unedited, acoustic recording of the ADOS examination, including the child’s vocal productions, but also those from the psychologist, as well as from the parents together with the ambient noise. In our recent publication, we showed that the duration of a preschooler’s vocal productions that could be identified by a diarization algorithm over the entire ADOS assessment was, on average, 3 minutes^48^, which corresponds to only 5-8% of the ADOS duration. The total duration of the child’s vocalizations was twice as lower for children who did not yet develop phrase speech. The low amount of child’s speech in our ADOS recorded with preschoolers suggests that, in addition to relying on the child’s voice features, our model considered a more broad acoustic signature of social interaction. Indeed, social interaction is a highly dynamic phenomenon; what each agent does or does not do will inevitably affect its quality. Bone and colleagues^36^ have shown that trained psychologist attune their prosody depending on the severity of autistic symptoms of the child while performing the ADOS with children aged 5-17 years old. The authors even reported that the psychologist’s prosodic cues better predicted the child’s autistic symptom severity than the prosody of the child himself. Additionally, an aspect of non-vocal information that might have helped the neural network to discriminate between children with and without autism resides in the presence of peculiar non-vocal acoustic features (i.e., ambient sound), such as any repetitive sound that would reflect an increased need for sensory stimulation in autistic children (e.g., repeatedly dropping objects or banging them, or perseverating on a musical toy…). The fact that we observed that sensory interests are the ADOS item that correlated the most with the diagnostic prediction based on the audio information would support the hypothesis that these non-vocal sounds played a role in the classification. The observation that autism can be accurately classified based solely on acoustic recordings of the ADOS assessment holds great potential for scaling autism screening. Indeed, in a context where the use of diarization pipelines to isolate speech from child and adult is not yet performing perfectly, and particularly when trying to discriminate between an adult female and a child’s voice^48^, the use of machine learning over audio recordings from segments of interactive play could not only prove to be a faster and more scalable option but also one that holds potential to reveal insightful information on the patterns of socio-communicative interaction between the child and an adult.

Given that the two neural networks achieved similar performance, the simple ensemble model did not outperform the results of any of the two networks taken separately. We would have expected each modality to capture different aspects of the broad autism spectrum symptoms uniquely. This was the case for some symptoms, such as sensory interests more related to audio prediction or the initiation of joint attention more related to video prediction. However, most autistic symptoms were relatively similarly predicted by both modalities, concordant with their complex and multimodal nature. Future work will be required to leverage better the different features that each neural network discriminates to combine them more efficiently. In the present work, we explored two ways of late fusion to achieve a combination that is either more specific (92.5%) or more sensitive (90%). In a context where early identification of children with autism is of utmost importance to provide them access to adequate intervention for supporting their development^49,50^, a highly sensitive screening tool would be most useful to minimize the number of children who would miss the opportunity of a further comprehensive assessment and opportunity for accessing intervention when needed. Our results strongly advocate for the feasibility of such a sensitive screening test exploiting naturalistic excerpts of the child’s social interactions in the natural environment. Unlike previous approaches^32,51^, the hereby solution does not require the child to wear any specific device, opening avenues for even more flexible and scalable screening solutions based on home video recordings. On the contrary, in circumstances when one might want to favor specificity over sensitivity, for instance, in the case of a primary practitioner concerned about a child’s development and who would like to confirm a prior suspicion of autism before referring the family for further in-depth assessment. As such, we suggest that using neural networks that combine audio and video information from naturalistic excerpts of social interaction is a promising direction for developing adaptive screening tools for low-cost, large-scale screening of autism spectrum disorder.

Advocating for combining information from different modalities should come as no surprise for clinicians involved in the diagnostic process. Indeed, actual best practices for diagnosis of autism recommend that the information obtained from the direct assessment of symptoms should be combined with detailed developmental history, cognitive and adaptive functioning measures, as well as genetic, medical, psychiatric, and behavioral conditions^52^. This inherently multi-modal process of clinician’s decision-making however remains yet to be more often exploited in the machine learning models aiming at the detection of signs of autism. Given the need for scalable tools that can be applied in less standardized contexts, the complementarity characteristic of multi-modal information could be an asset, especially in the contexts of lower validity^47,53^. In other domains, such as brain imaging, combining different modalities has shown to represent a promising manner to increase classification accuracy^54–56^. A recent study that used acoustic analyses combined with a computer vision approach applied in the context of the paradigm of response to name has achieved 92% consistency with clinical ground-truth ratings^57^. Recently a large, multi-site study implemented a multi-modal, app-based approach to screening for autism signs that significantly outperformed the traditional parent-report screening questionnaires^58^. The multimodality approach shows tremendous potential not only in early detection but also in the successful stratification of subtypes of autism. Indeed, a recent study using a wide array of patient health data, such as electronic health records, healthcare claims, familial whole-exome sequences, and neurodevelopmental expression patterns, together with state-of-the-art AI methodology, managed to successfully isolate a subtype of autism characterized by dyslipidemia^59^. Future studies should be conducted to explore whether multimodal assessments automatically extracted from simple audio-video recordings of social interactions hold a similar potential for identifying reliable and meaningful autism subgroups, which would represent a critical step toward precision medicine for autism.

The constant growth seen in the development of novel technologies and their scalability holds a true potential to radically change the landscape of psychiatry in the years to come. The ability to efficiently integrate meaningful features into a coherent pattern, characterizing the thought process of highly skilled clinicians, is now possible on an ever-increasing scale. In the case of a disorder as heterogeneous as autism, the possibility to efficiently extract and combine huge amounts of meaningful features is a critical step towards much-needed more reliable screening, diagnosis, and progress monitoring.

## Methods

### Ethics

The study and protocols conducted in this research were approved by the Ethics Committee of the Faculty of Medicine at the University of Geneva, Switzerland. The methods employed in this study strictly adhered to the relevant guidelines and regulations set forth by the University of Geneva. Informed written consent was obtained from a parent and/or legal guardian for all children participating in the study. Children diagnosed with Autism Spectrum Disorder (ASD) met the clinical criteria outlined in DSM-5^1^, and their diagnosis was further supported by employing gold standard diagnostic assessments (refer to the Supplementary section for more details). Typically developing (TD) children underwent screening to ensure the absence of any known neurological or psychiatric conditions and no history of ASD in any first-degree relatives. Informed consent was obtained from the subject in Figure 1, their legal guardian and the examiner for the publication of identifying information/images in an online open-access publication.

### Participants

Participants were enrolled as part of the Geneva Autism Cohort, a longitudinal project aiming to better understand the early development of children with autism spectrum disorder (ASD)^60–62^. The dataset is composed of 80 children with ASD (2.68 ± 0.89 years old; 48 males, 32 females) as well as 80 typically developing (TD) children matched for age (2.46 ± 0.98 years old) and gender (48 males, 32 females). Children were included in the ASD group following expert clinical diagnosis using the DSM-5 criteria^63^, and upon confirmation by gold standard diagnostic assessment (see below). TD children were screened for the presence of any known neurological or psychiatric disorder and had no known first-degree relative with ASD.

All children underwent the Autism Diagnosis Observation Schedule (ADOS)^3,4^, which was video recorded. All autistic children included in the current study had ADOS scores superior to the threshold for autism. As in our previous work^25^ for the measure of the cognitive level, we used Best Estimate Intellectual Quotient^64,65^, a measure that amalgamates the most pertinent cognitive assessment measures for each child. In addition, adaptive functioning was estimated using the Vineland Adaptive Behavior Scales, second edition (VABS-II)^66^.

A detailed description of the sample characteristics can be found in Supplementary Table S1). All participants or their legal guardians gave written permission following relevant regulations and guidelines set forth by the Faculty of Medicine Ethics Committee of Geneva University, Switzerland.

### Video and Audio Datasets

To obtain a training and testing sample we used a 50-50 split of the sample of 160 individuals. Both training and testing samples included 40 children with Autism Spectrum Disorder (ASD) and 40 typically-developing children matched for age, sex, and the ADOS module that was used (see Supplementary Table S1).

### Video Neural Network

For the analyses of the visual features from the ADOS videos, we adopted an approach similar to the one used in previous work^25^. We first proceeded to dimensionality reduction by applying pose estimation^26^ on the ADOS session to isolate body movements and gesture-based interactions. The extracted skeletal key points were plotted over a plain black background preserving a visual gist of the social interaction between the examiner and the child. The reduced-dimensionality videos were further down-scaled into 320×240 resolution – similar to UCF-101^67^. Videos were then split into 5-second segments to avoid over-fitting (see Supplementary Table S1). The resulting processed dataset consisted of 43633 five-second video clips in training (Average Video Length: 45.45 min; Total Duration: 60.601 hours) and 44796 video clips in the testing set (Average Video Length: 46.66 min; Total Duration: 62.22 hours) for the video classifier.

Our model architecture takes the 5-second video clips as inputs. It uses an ImageNet pre-trained VGG16 convolutional neural network to extract high-dimensional features from each frame of the inputted segment. This was done by removing the fully connected dense layers in the VGG16 architecture. These high dimensional features were input to 512 hidden layered LSTM RNN^68^ followed by 512 fully connected Dense Layers. We trained the neural network using an 80-20 training validation split. We carried out hyperparameter tuning and observed the best configuration for the VGG16 LSTM RNN to be at 625 batch size and 100 epochs. To reduce overfitting, we introduced a dropout of 0.5 followed by a softmax classification layer^69^ that gives an output of 2 classes ASD and TD. To compute the loss we use categorical cross entropy^70^ with a rmsprop optimizer^71^.

We then used the trained model to carry out predictions over the pre-processed 5-second video clips from the testing set to obtain segment-wise predictions (see Supplementary Figure S3). In the next step, we only retain the predictions that have a confidence of more than 90% which on average removes 29% of the segment-wise predictions across the testing dataset, yet at a lower confidence by the video neural network.

This approach of retaining only segments predicted with more than 90% confidence ensures prediction performance stability.

Finally, the segment-wise predictions are further averaged for the duration of the entire video. When the averaged prediction confidence for ASD is above 50%, the video is assigned to the ASD class, otherwise the predicted class is TD (see Figure 1).

### Audio Neural Network

Upon extraction of the audio track from the ADOS recording, audio files were converted to a 2-channel, 44kHZ sample rate, and 192k bitrate format. This standardized format was chosen to homogenize the audio format from different microphones used for ADOS recordings and match the parameters of audio samples in the Urban8K dataset^72^, a dataset used to benchmark several audio neural networks^73^. We then split the audio files into segments of 10 seconds each, as this duration proved optimal for training the model (see Supplementary Table S3). The resulting dataset consisted of 22^*′*^466 10-second audio clips for the training (Average Length: 46.804 min; Total Duration: 62.41 hours) and 22430 clips for the testing set (Average Length: 46.73 min; Total Duration: 62.3 hours).

The neural network architecture is inspired by the 1-dimensional Convolutional Neural Network (1D-CNN)^74^ with different audio features extracted from audio clips added to enhance classification^75^. These features include mean Mel Frequency Cepstral Coefficients (MFCCs)^41^, Mean Chromagram^44^, Mean Mel Spectrogram^40^, Mean Spectral Contrast^43^ and Mean Tonal Centroid^42^. The features are concatenated into a one-dimensional vector by taking a mean along the time axis for each of the 10-second segments. This vector is then used as an input for convolution and classification. In the CNN, the vector is passed through two 1-dimensional convolutional layers followed by a Maxpooling layer^76^ and another 1-dimensional convolutional layer. To decrease overfitting a dropout of 0.5 is used followed by flattening^77^ of the output. This is then passed through 512 fully connected dense layers and a softmax classification layer to get the final output of two classes, ASD and TD. We used categorical cross-entropy with an Adam optimizer^78^ to compute the loss. A procedure similar to the video neural network was applied to train the audio CNN. The training data was divided into an 80-20 split (80% of the dataset being used for training and 20% for validation). We found that the optimal set of MFCC features to input into the CNN is 150. We proceeded to hyperparameter tuning and established 512 as the batch size and 30 epochs respectively (see Figure 2).

After training the model, we made predictions over 80 audio files of the testing set. The pre-processing steps were consistent with the training data in which each audio file was converted into an optimized format and splitting the audio files into 10-second segments. We then used the trained model to carry out segment-wise predictions over these audio clips using CNN. The predictions for each audio clip are aggregated by taking the average confidence across all segments subject-wise. The median of the average confidence decides the overall audio file classification as ASD or TD.

### Combining Video and Audio Neural Network

Different approaches were investigated regarding the combination between the audio and the video neural networks for classification algorithms with the highest 1) sensitivity, 2) specificity, and 3) classification accuracy. To obtain a classifier with the highest sensitivity, we created a model where the final prediction would be ASD if either neural network resulted in a classification of ASD (that we denoted *OR model*). By doing so, we expected that our classifier would yield less false negatives diagnoses (thus being more sensitive), at the cost of lower specificity. We also explored the possibility to get a more specific classifier, for which the final prediction would be ASD only if both neural networks resulted in a classification of ASD (that we denoted *AND model*). This classifier is expected to yield less false positives (thus being more specific), at the cost of sensitivity. Finally, we explored different combinations of the two neural networks to optimize the accuracy. For this purpose, we manually applied different thresholds to combine both predictions, depending on the confidence in the prediction of each network, to yield an *ensemble* network that would have the highest accuracy.

### Analysis of Video-Audio Neural Network Predictions with ADOS scales

To test the clinical relevance of the machine learning algorithm, we independently verified that our results align with the standardized clinical measures of autistic symptoms, as measured with the 27 individual ADOS items (see Supplementary Table S3).). Specifically, we examined whether the accuracy in identifying ASD by either modality (i.e., audio *vs* video) depended upon the child’s profile. To that end, we tested how the ASD prediction from each of our neural networks relates to the ADOS items assessed on a scale ranging from 0 (least evidence of autistic signs) to 3 (marked presence of autistic signs).

Of note, the ADOS manual prescribes coding specific behavioral features that may have clinical relevance but are not of the autistic quality with codes that are not comprised between 0 and 3. For instance, a code of 7 is used when the behavior shows an abnormality, but its quality is not obviously autistic. A code of 8 is given when the behavioral level necessary for assessing the symptom is not attained at the time of the assessment (e.g. the child’s language level is too low to assess the stereotyped quality). Finally, a score of 9 is given when the specific social press was not correctly administered. According to the ADOS, coding guidelines scores 7-9 are recoded as 0 in the final diagnosis algorithm to not bias the final score by measures that do not reflect autistic symptomatology. In the present study, to distinguish these cases from clearly typical behavior that is denoted with a 0, we recoded scores 7-9 as -1.

For each modality (audio & video), we then used a random regression forest model^45^ with the 26 items of the ADOS as predictors and the confidence of the neural network as the dependent variable. We calculated how much the mean square error (MSE) increased when a specific ADOS item was omitted from our model to determine each item’s importance. The higher the percentage increase in MSE (%IncMSE), the higher the contribution to the prediction^79^.

For each modality, we delved deeper into a more detailed multi-way importance estimation^80^ by using %IncMSE (mse_increase), increase in node purity (node_purity_increase), and p-value to hierarchize the importance of each ADOS item in predicting neural network confidence. An increase in node purity translates into higher predictor importance. The predictor is considered significant if the p-value is less than 0.05.

Finally, we compared the strength of the association between the ADOS items and the prediction confidence between the two modalities (audio *vs* video), considered separately. As the %IncMSE could not be directly compared from one Random Forest Regression Model to another, we calibrated another Random Forest Regression Model wherein the averaged confidence scores from both of the neural networks were used as the value to be predicted, and items from ADOS-2 were used as predictors. The %IncMSE values obtained from the *average confidence* Random Regression Forest Model were then used to normalize the %IncMSE values of the audio and video neural networks by subtracting and dividing their %IncMSE values with the %IncMSE values obtained from the average confidence model. We thus obtained standardized %IncMSE values for both the audio and the video neural network Random Regression Forest Models. These standardized %IncMSE values were then used to create a linear regression model with residual scores for the audio and video cases. These residual scores were then plotted against each other to visualize the relationship between different ADOS-2 items and the two neural networks (see Figure 5).

## Data Availability

Code used for developing video and audio neural network is available by the corresponding author upon request. The ADOS clinical examination recordings from which the video and audio data were extracted represent sensitive data and thus cannot be shared.

## Acknowledgements

We are grateful to all the families involved in this study and the psychologists and colleagues who contributed to the data collection.

This study was funded by the Sinergia Grant for Digital Phenotyping of Autism Spectrum Disorders in Children (Grant Number: 202235), National Centre of Competence in Research (NCCR) Synapsy, by the Swiss National Science Foundation (SNF, Grant Number: 51NF40_185897), by SNF grants to M.S. (#163859 and #190084), as well as by funds from the Private Foundation of the HUG, by a UNIGE COINF2018 equipment grant and by the Fondation Pôle Autisme (https://www.pole-autisme.ch). We are also very grateful to the Alexis for Autism initiative (https://www.alexisforautism.com) for supporting this research.

## Author contributions statement

SN, NK, TM, and MS conceptualized the research study. NK was responsible for data curation and ran the pose estimation on ADOS videos. SN carried out further pre-processing of the data and developed the audio and video neural network models. SN carried out analysis using random regression forest models. TM carried out analysis using regression modeling. SN and NK wrote the initial manuscript draft, which all authors edited.

