## Supplementary Material PDF for "Video-Audio Neural Network Ensemble For Comprehensive Screening Of Autism Spectrum Disorder in Young Children"

### Supplementary Information

#### Dataset

The Table S1 summarizes the clinical characteristics of the Training and Testing samples. The sample preparation was constrained by fewer available ADOS recordings from typically developing (TD) children in our larger longitudinal cohort based in Geneva. The available TD sample of TD children at the time of this study included 80 children (32F) and was used to select an age and gender-matched sample of 80 children with ASD. This global sample of 160 videos was split into two closely matched Training and Testing sets. Children with ASD in both samples showed a moderate to high level of autistic symptoms, as illustrated by their average ADOS-CSS of 7.43 and 7.85, respectively. No significant differences in all used clinical measures were found between the Training and Testing samples (all p values of group differences were superior to 0.05).

| **Measures** | **Training set (*n* = 80)** | | **Testing set (*n* = 80)** | | ***p* value** |
|---|---|---|---|---|---|
| **Distribution** | ASD (*n* = 40) | TD (*n* = 40) | ASD (*n* = 40) | TD (*n* = 40) | |
| **Gender** | 32F / 48M | | 32F / 48M | | |
| **Modules (MT / M1 / M2)** | 32 / 34 / 14 | | 35 / 30 / 15 | | |
| | *Mean(SD)* | *Mean(SD)* | *Mean(SD)* | *Mean(SD)* | |
| Age | 2.79±0.853 | 2.52±0.891 | 2.61±0.974 | 2.40±1.09 | 0.306[a] |
| Total Symptom Severity Score (CSS) | 7.43±1.77 | 1.0±0.0 | 7.85±1.78 | 1.08±0.267 | 0.571[b] |
| Social Affect (SA-CSS) | 6.55±2.03 | 1.05±0.221 | 7.15±2.02 | 1.13±0.404 | 0.553[b] |
| Repetitive Behaviors & Restricted Interests (RRB CSS) | 8.85±1.53 | 1.6±1.45 | 8.43±1.69 | 2.58±2.07 | 0.758[b] |
| Best Estimate IQ | 78.8±25.6 | 120±18.7 | 74.7±25.6 | 112±15.5 | 0.279[a] |
| VABS-II Adaptive Behavior | 78.1±9.32 | 101±8.86 | 78.4±10.5 | 102±7.38 | 0.689[a] |
| VABS-II Communication | 75.1±14.4 | 103±9.35 | 75.4±13.8 | 104±10.3 | 0.730[a] |
| VABS-II Daily Living Skills | 82.6±10.9 | 102±9.30 | 81.3±12.3 | 102±8.57 | 0.951[a] |
| VABS-II Socialization | 77.6±7.91 | 100±9.26 | 80.3±9.59 | 101±6.37 | 0.333[a] |
| VABS-II Motor Skills | 89.3±9.81 | 101±8.63 | 87.9±12.0 | 101±10.0 | 0.933[a] |

*Note*. Normality was checked for the data using Kolmogorov-Smirnov Test. P-values are obtained using a t-test for two independent sample[a] or nonparametric Mann-Whitney[b] tests of differences between the Training & Testing set.

**Table S1.** Description of the Training and Testing sets used in the present study.

#### Preprocessing of dataset

Upon the creation of the balanced training and testing set, we carried out pre-processing as discussed in the Methods section to obtain 5-second and 10-second segments that were used for training the video and audio neural networks respectively. The details about the segments obtained after carrying out pre-processing, the mean length of each subject's examination recording, and the total time of recording in the training and testing sets can be observed in the Table S2. As can be observed, we were able to obtain a total of 43,633 segments for the video training set, and 44,796 segments for the video testing set. 22,466 segments for the audio training set, and 22,430 segments for the audio testing set. The mean duration for the videos in the training set was 45.451 minutes and 46.663 min for the testing set. In the case of audio, the mean duration of the audio recordings was 46.804 minutes and 46.729 minutes.

| **Parameter** | **Video Training set** | **Video Testing set** | **Audio Training set** | **Audio Testing set** |
|---|---|---|---|---|
| No. of Videos | 80 | 80 | 80 | 80 |
| No. of segments | 43633 | 44796 | 22466 | 22430 |
| Total Duration (hours) | 60.601 | 62.217 | 62.406 | 62.306 |
| Average Video Length (min) | 45.451 | 46.663 | 46.804 | 46.729 |

**Table S2.** The distribution of audio and video datasets extracted from the ADOS clinical examination recordings for creating Training and Testing sets. These sets were then inputted into the audio and video neural networks respectively carry out training and later on prediction.

#### Feature Extraction

While carrying out the development of the audio and video neural networks, we also carried out visualization of the features that were used as input to the neural network systems. The process of visualizing features was majorly carried out to ensure that null or empty features were not used as input for the neural network to train upon.

**Video Neural Network Features:** We first carried out a visualization of features that were extracted using the VGG16 CNN from the frames of the 5-second videos and used as input for the LSTM units. We implemented GradCam[81] to generate saliency maps using gradient-based localization over each of the frames extracted from the 5-second video clips (See Figure S1 A.). These saliency maps or heatmaps generated through GradCam represented significant regions of each of the frames that were used as input features for the carrying out training and prediction using the LSTM RNN (See Figure 1). As can be observed in the visualization, the skeletons of the examiner (on the right) and child (on the left) were significant and were utilized by the LSTM RNN for training and prediction. The segment number of the video clip (Segment), prediction (Pred), truth(Truth), the confidence of ASD prediction (prob_ASD), and confidence of TD prediction (prob_TD) can be seen at the top of the illustration.

**Audio Neural Network Features:** To carry out training and prediction using the convolutional neural network over the audio samples, we extracted 5 different types of features from the 10-second audio clips namely Mel spectrogram[40], MFCCs[41], Tonal Centroids (Tonnetz)[42], Chromagram[44] and Spectral Contrast[43].

### Audio Neural Network Training

We tested out the audio neural network over several different hyper-parameters but observed the best batch size to be at 512 with 30 epochs at an 80-20 training validation split for the audio neural network training. We extracted 150 Mel Frequency Cepstral Coefficients from each of the 10-second segments that we extracted from the original audio files and used a 1-dimensional Convolutional Neural Network Architecture (See Figure 2) in order to carry out training. We split 80% of the original training data into newer training data and 20% into validation data.

We then used the training model to carry out predictions over the testing data to test the accuracy of the trained audio neural network model. The training history and confusion matrix for the predictions made over 80 videos in the testing set can be seen in Figure S2.

### Video Neural Network Training

We carried out balancing of our dataset (as discussed in Methods Section) by deleting segments that were less than 10 kilobytes in size to have less noise when training the neural network. Since we observed that the VGG16 LSTM RNN showed good performance for training and prediction with our OpenPose normalized dataset, we used the same neural network architecture for creating the video neural network classifier.

Furthermore, we used a filtering condition during the process of prediction wherein only predictions with more than 90% confidence over the video segments were taken into account when aggregating the overall prediction for a given subject's video. Using this pipeline we were able to achieve an accuracy of 80% (See Figure 1). The training plot for the VGG16 LSTM RNN and confusion matrix for the predictions made by the neural network can be observed in Figure S3.

### Audio-Video Ensemble Neural Network

We used different ways to fuse the audio and video neural networks to create an ensemble (as discussed in Methods Section). The systems that gave us the best result used the condition where if the confidence of the video neural network prediction is less than 60% we used the confidence from the audio neural network (See Figure 3). Using this system we were able to achieve an accuracy of 82.5%. We also carried out testing the sensitivity of the two neural network systems. In this test, we first took the final prediction as ASD if either one of the audio or video neural network predicts it as ASD (OR model). In the second test, we only took the final prediction as ASD if both neural network predictions were ASD (AND model). Using this we were able to obtain a confusion matrix. The confusion matrix for the AND and OR test as well as the ensemble neural network can be seen in Figure S4.

### Multiple layers for screening ASD

One of the interesting aspects of this study for us was also to identify the number of times, both the neural networks failed to correctly predict a subject as it represents how effective the multi-layered/multi-modality-based approach is in the screening of ASD over a larger scale. We found out from our results that both neural networks incorrectly predicted only 8.75% of subjects among the 80 subjects that were present in the testing sample. This provided us with reassurance that our system proves to be very robust and sensitive with a very low number of false negatives. The confusion matrix for the number of videos incorrectly predicted by both audio and video neural networks can be observed in Figure S5

| Behaviors | %IncMSE Video | %IncMSE Audio |
| --- | --- | --- |
| **A. Communication** | | |
| Frequency of Spontaneous Vocalization Directed to Others | -0.122 | -4.216 |
| Intonation of Vocalizations or Verbalizations | 0.852 | -3.198 |
| Immediate Echolalia | 1.327 | 2.117 |
| Stereotyped/Idiosyncratic Use of Words or Phrases | 2.809 | -3.757 |
| Use of Another's Body | 0.984 | -6.136 |
| Pointing | 5.836 | 10.232 |
| Gestures | 5.122 | -3.272 |
| **B. Reciprocal Social Interaction** | | |
| Unusual Eye Contact | 7.306 | 6.072 |
| Facial Expressions Directed to Others | 6.066 | 5.188 |
| Integration of Gaze and Other Behaviors during Social Overtures | 4.079 | 1.961 |
| Shared Enjoyment in Interaction | 4.614 | 3.654 |
| Response to Name | 3.468 | 9.414 |
| Requesting | 4.782 | 2.509 |
| Showing | -4.444 | 14.770 |
| Giving | 4.934 | -4.938 |
| Spontaneous Initiation of Joint Attention | 9.645 | -0.654 |
| Response to Joint Attention | -1.743 | -2.105 |
| Quality of Social Overtures | 9.568 | 9.005 |
| Amount of Social Overtures/Maintenance of Attention: Examiner | 4.459 | 7.762 |
| Quality of Social Response | -0.229 | -1.436 |
| Quantity of Social Communication | 0.730 | -4.404 |
| Overall Quality of Rapport | 6.091 | 0.700 |
| **C. Play** | | |
| Functional Play with Objects | 3.077 | 0.877 |
| Imagination / Creativity | 5.600 | 0.649 |
| **D. Stereotyped Behaviors and Restricted Interests** | | |
| Unusual Sensory Interest in Play Material/Person | 6.172 | 15.832 |
| Hand and Finger and Other Complex Mannerisms | 2.618 | 13.261 |
| Unusually Repetitive Interests or Stereotyped Behaviors | -0.399 | 2.626 |

**Table S3.** Selected items from the gold-standard diagnostic assessment, the ADOS-G[3] and ADOS-2[4] which were used to create random regression forest model to estimate their importance with respect to audio and video neural network's prediction confidence. The different columns correspond to the behavior and percentage increase in mean square error (%IncMSE) for each of the items with respect to video and audio neural network's confidence respectively extracted through the random regression forest model using the items as predictors for prediction the neural network's confidence.

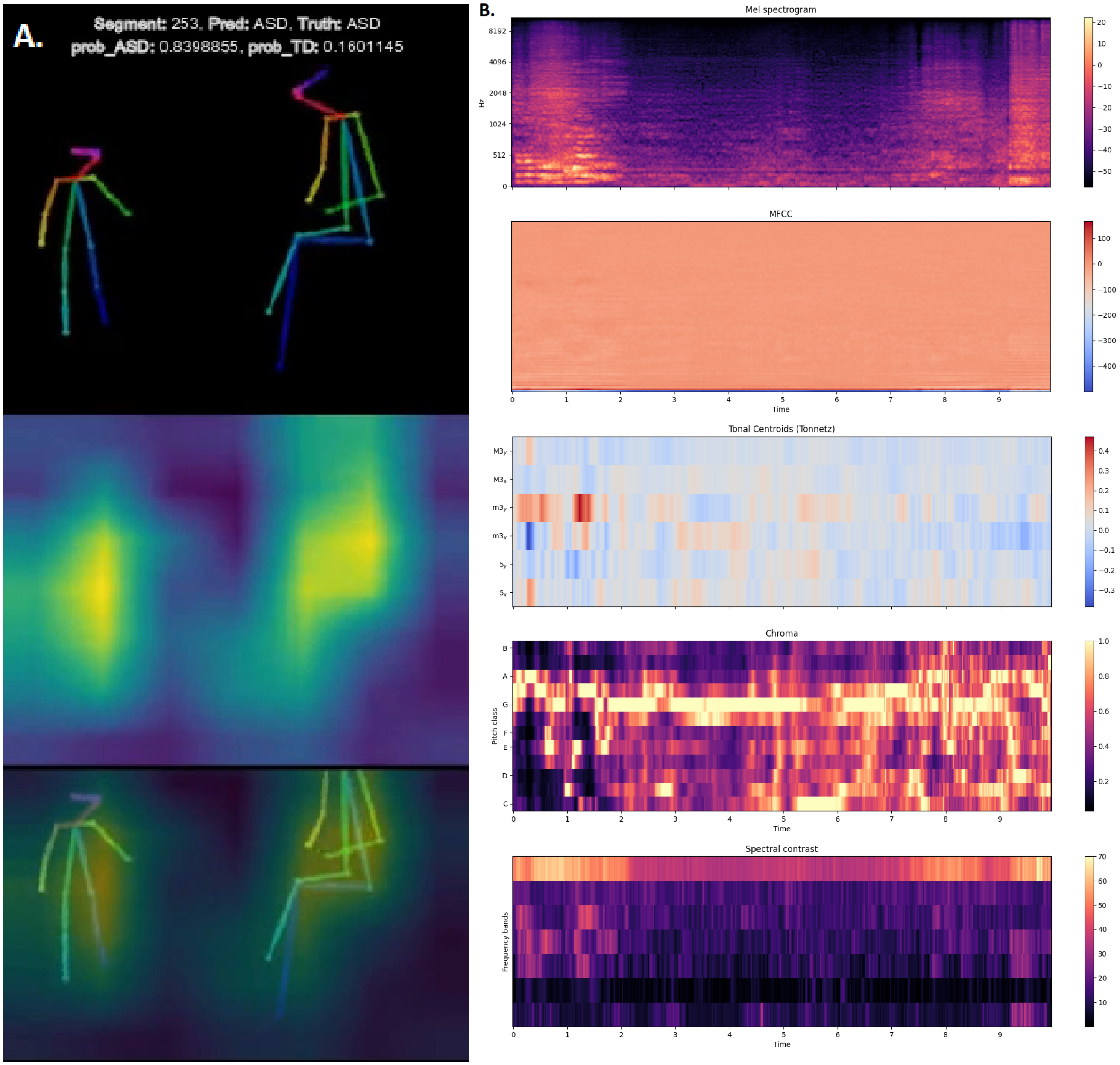


**Figure S1.** **A.** Implementation of VGG16 GradCam which generates a saliency or heat map showcasing significant regions in each frame that were used as feature inputs to the LSTM RNN for training the video neural network**B.** The 5 features extracted from the 10-second audio clips (Mel Spectrogram, MFCCs, Tonal Centroids, Chromagram and Spectral Contrast) that were flattened into a vector and used as input to the CNN used for carrying out training and classification of audio samples.

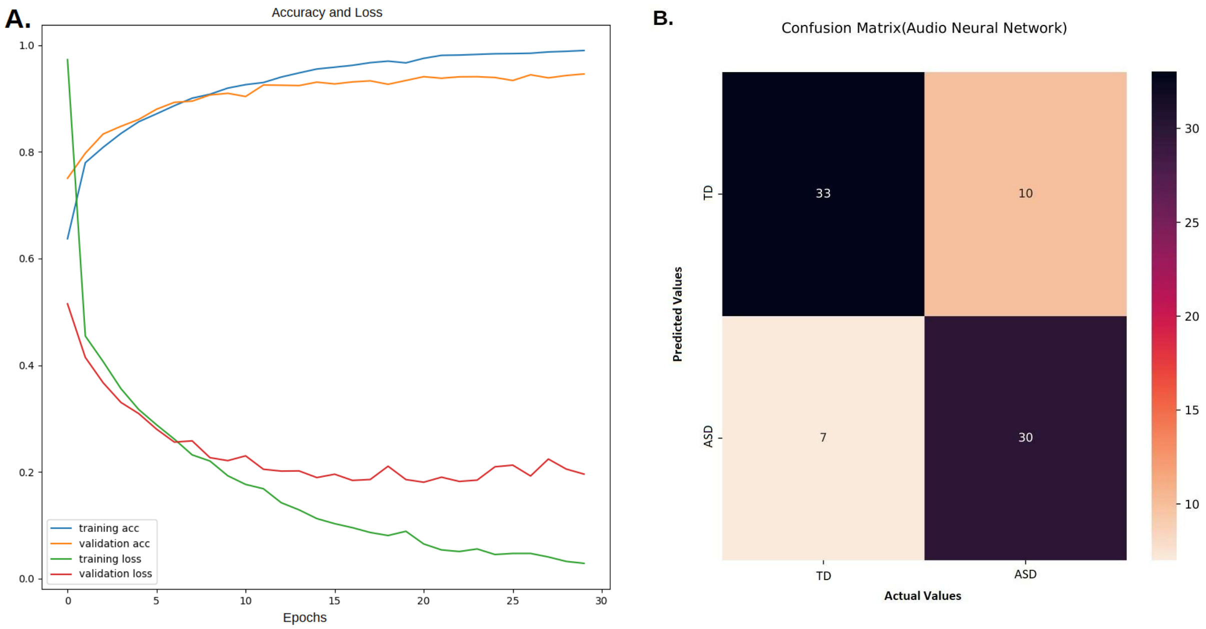


**Figure S2.** **A.** represents the Training Accuracy, Training Loss, Validation Accuracy, and Validation loss with 80 balanced training audio dataset at 80-20 split, 512 Batch Size, 2-channel, 44100 Hz Sampling rate, 192k bitrate, 10-second segments and 30 Epochs for 1-dimensional Convolutional Neural Network for audio classification. **B.** is a visual representation of the confusion matrix obtained from the audio neural network predictions over 80 pre-processed audio files that were extracted from the testing sample

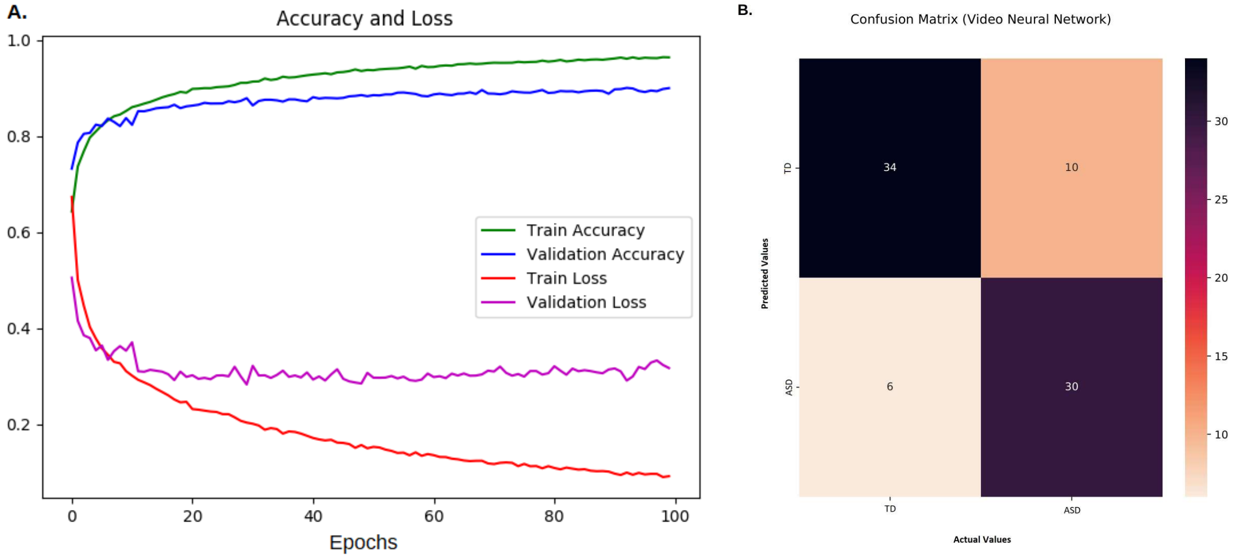


**Figure S3.** Training Accuracy, Training Loss, Validation Accuracy and Validation loss with 80 balanced training audio dataset at 80-20 split, 625 Batch Size, 320X240 resolution, 5-second segments and 100 Epochs for VGG16 LSTM RNN for video classification

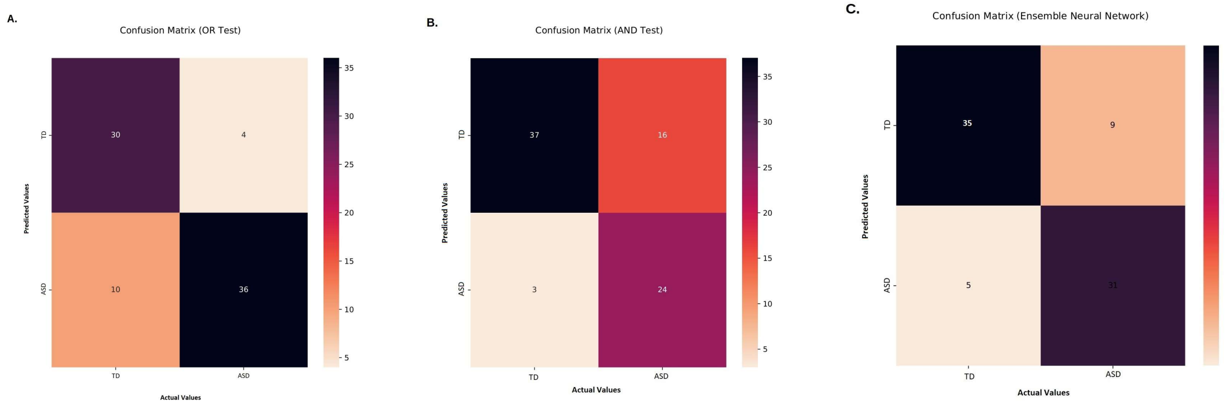


**Figure S4.** Confusion matrices obtained from the **A.** OR model, **B.** AND model and **C.** video audio neural network ensemble. The figures represent false positives, false negatives, true positives, and true negatives which were used to obtain accuracy, precision recall, specificity, negative predicted values and F1-score

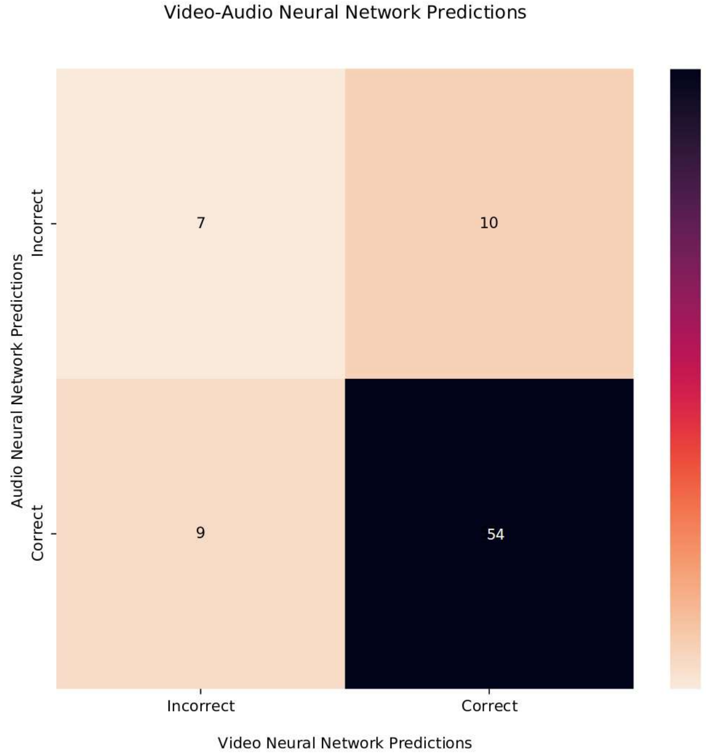


**Figure S5.** Confusion Matrix representing predictions made by audio and video neural networks over the 80 video-audio testing set wherein video neural network achieved an accuracy of 80% (64/80 patients correctly predicted) and audio neural network achieved an accuracy of 78.75% (63/80 patients correctly identified), only 8.75% (7/80 patients) were incorrectly identified by both audio and video neural networks indicated the chance of failure of both neural networks in predicting ASD is 8.75%
